# Integrative analysis reveals a macrophage-predominant, immunosuppressive immune microenvironment and subtype-specific therapeutic vulnerabilities in advanced salivary gland cancer

**DOI:** 10.1101/2024.06.11.24308538

**Authors:** Erika Zuljan, Benjamin von der Emde, Iris Piwonski, Ana Cristina Afonseca Pestana, Konrad Klinghammer, Andreas Mock, Peter Horak, Christoph Heining, Frederick Klauschen, Ina Pretzell, Melanie Boerries, Christian H Brandts, Simon Kreutzfeldt, Maria-Veronica Teleanu, Daniel Hübschmann, Luc G T Morris, Ulrich Keller, Hanno Glimm, Stefan Fröhling, Sebastian Ochsenreither, Ulrich Keilholz, Eric Blanc, Dieter Beule, Damian T Rieke

**Affiliations:** Core Unit Bioinformatics, Berlin Institute of Health at Charité–Universitätsmedizin Berlin, Charitéplatz 1, Berlin, Germany; Department of Hematology, Oncology and Cancer Immunology, Campus Benjamin Franklin, Charité–Universitätsmedizin Berlin, Corporate Member of Freie Universität Berlin and Humboldt-Universität zu Berlin, Berlin, Germany; Department of Pathology, Charité-Universitätsmedizin Berlin, Corporate Member of Freie Universität Berlin and Humboldt-Universität zu Berlin, 10117 Berlin, Germany; Comprehensive Cancer Center, Charité - Universitätsmedizin Berlin, Corporate Member of Freie Universität Berlin and Humboldt-Universität zu Berlin, Chariteplatz 1, 10117, Berlin, Germany; German Cancer Consortium (DKTK) and German Cancer Research Center (DKFZ), partner sites Berlin, Dresden, Essen/Düsseldorf, Freiburg, Heidelberg and München; Institute of Pathology, Ludwig-Maximilians-University Munich, Thalkirchner Str. 36, 80337 München, Germany; Division of Translational Medical Oncology, German Cancer Research Center (DKFZ), Heidelberg, Germany; National Center for Tumor Diseases (NCT), NCT Heidelberg, a partnership between DKFZ and Heidelberg University Hospital, Heidelberg, Germany; Department for Translational Medical Oncology, National Center for Tumor Diseases Dresden (NCT/UCC), a partnership between DKFZ, Faculty of Medicine and University Hospital Carl Gustav Carus, TUD Dresden University of Technology, and Helmholtz-Zentrum Dresden - Rossendorf (HZDR), Germany; Translational Medical Oncology, Faculty of Medicine and University Hospital Carl Gustav Carus, TUD Dresden University of Technology, Dresden, Germany; West German Cancer Center, University Hospital Essen, Essen, Germany; Institute of Medical Bioinformatics and Systems Medicine, Medical Center-University of Freiburg, Faculty of Medicine, University of Freiburg, Freiburg, Germany; Goethe University Frankfurt, University Hospital, Department of Medicine, Hematology/Oncology & University Cancer Center Frankfurt (UCT), Theodor-Stern-Kai 7, 60590 Frankfurt, Germany; Department of Surgery, Memorial Sloan Kettering Cancer Center, New York, New York, USA; Institute of Human Genetics, Heidelberg University, Heidelberg, Germany

**Author notes:** equal contribution (Eric Blanc, Dieter Beule, Damian T Rieke). Correspondence: Damian T. Rieke, Charité Comprehensive Cancer Center, Charitéplatz 1, 10117 Berlin.

**Keywords:** Salivary Gland Cancer, Tumor Immune Microenvironment, Immune Therapy, Immune Checkpoints, Macrophage, T-cells

## Abstract

**Background:** Salivary gland cancers (SGC) are rare and heterogeneous malignant tumors. Advanced SGC lack established treatment options and show poor response to immunotherapy. Here, an integrative multi-omics analysis in a large cohort of advanced SGC revealed insights into the tumor immune microenvironment (TIM) and distinct mechanisms of immune evasion.

**Methods:** A total of 104 patients with recurrent/metastatic SGC from the DKTK MASTER program were included in this study. Whole-exome or whole-genome sequencing and RNA-sequencing was performed on fresh frozen tumor tissue. The tumor immune microenvironment was analyzed using CIBERSORT deconvolution analysis and immune gene expression scores in bulk RNA-sequencing data. Single-nuclei sequencing and immunohistochemistry analyses were performed in selected samples. Results were validated in bulk RNA-sequencing data of a previously published independent dataset.

**Results:** Bulk transcriptome analysis revealed an immune-deserted TIM in the majority of advanced SGC samples. Immune exclusion was most prominent in adenoid cystic carcinoma (ACC) subgroup 1 exhibiting a downregulation of the antigen processing machinery. Only a small subset of advanced SGC, including few adenoid cystic carcinoma, exhibited T-cell inflammation, which was correlated with tumor mutational burden in Non-ACC samples. Subtype specific expression of immune checkpoints as well as cancer testis antigens were identified with prominent expression of VTCN1 in luminal cells within ACC. Single-cell RNA-sequencing and bulk RNA-seq deconvolution analysis validated immune cell exclusion and revealed a TIM that was dominated by M2 macrophages across SGC subtypes. Among evaluable patients treated with immune checkpoint inhibitors, a high T-cell to macrophage ratio was associated with clinical benefit.

**Conclusions:** These data support biomarker-based development of immune-checkpoint inhibition and the development of novel immune-checkpoint inhibitors and cellular therapies in SGC.

**Trial Registration:** Retrospectively registered, NCT05852522

## Background

Salivary gland cancers (SGC) are a rare and heterogeneous group of malignancies that arise from major and minor salivary glands. SGC account for 5% of all head and neck cancers and comprise more than 20 different histologies ^1^. The prognosis is poor in the recurrent and metastatic setting, with a median OS of 15 months after appearance of distant metastasis ^2^. No approved therapies for advanced SGC exist. Histological subtypes are often not adequately represented in available clinical trials with the exception of adenoid cystic carcinoma (ACC). ACC is among the most common malignant subtypes, defined by a recurrent *MYB-NFIB* gene fusion and often presents with slow tumor growth and poor response to systemic therapy ^3^. Yet, different molecular and clinical subsets of ACC have been described ^4^. In contrast, non-ACC tumors typically show an aggressive clinical course and often harbor targetable molecular alterations ^5^.

Immune checkpoint inhibition yielded low response rates in clinical trials in advanced salivary gland cancer. In a study of pembrolizumab in PD-L1 positive salivary gland cancer, 26 patients were enrolled and three objective responses (all PR) could be reported in adenocarcinoma (n=2) and high-grade serous carcinoma (n=1) ^6^. Similarly, responses were observed in several histologies, including ACC, in another phase 2 trial of pembrolizumab in 109 patients with pretreated SGC for an objective response rate of 4.6% ^7^. In this trial, a higher response rate (10.7%) was noted in PD-L1 positive disease ^7^. A comparable response rate of 4.2% was reported in a retrospective multicenter analysis of nivolumab in 24 patients with SGC ^8^. In this analysis, a response was observed in a patient with salivary duct carcinoma ^8^. The combination of nivolumab and ipilimumab was tested in separate cohorts of 32 patients each with adenoid cystic and non-adenoid cystic carcinoma ^9^. In this trial, the primary efficacy endpoint of at least 4 objective responses was met in the non-ACC (ORR 16%) but not in the ACC cohort (ORR 6%) ^9^. Similarly, no objective responses were noted in 20 adenoid cystic carcinoma patients randomized 1:1 to pembrolizumab with or without radiotherapy ^10^. These data show an overall limited efficacy of immune checkpoint inhibitors in advanced SGC ^6,9–11^. Among these prospective trials, no clear subtypes and predictive biomarker for the use of immune checkpoint inhibitors could be identified. The use of immunotherapy outside of clinical trials is therefore not routinely recommended in SGC and predictive biomarkers and novel treatment strategies are urgently required ^12^. Available data on the tumor immune microenvironment in SGC show subgroup-specific differences with an immune-excluded microenvironment in adenoid cystic carcinoma but only limited data are available in the recurrent and/or metastatic setting ^13–15^. Here, we provide a multi-omics analysis of the tumor immune microenvironment in a cohort of recurrent and/or metastatic salivary gland cancers, thus representing a potential intention-to-treat cohort for systemic therapies. In doing so, we are able to describe relevant immune-infiltration in a subset of tumors, including a limited number of adenoid cystic carcinomas. A higher tumor mutational burden is observed in inflammation-high SGC. We demonstrate a macrophage-predominant microenvironment in all SGC entities. Analysis of patients treated with immune checkpoint inhibitors shows a clinical benefit in individual cases with a T-cell predominant TIM. Additional analyses of potential novel targets for immunotherapy reveals significant overexpression of *VTCN1* (V-Set Domain Containing T Cell Activation Inhibitor 1) **in** adenoid cystic carcinoma and expression of cancer-testis antigens in a majority of SGC. Taken together, these results provide a comprehensive insight into the tumor immune microenvironment in advanced SGC.

## MATERIAL AND METHODS

### Participant characteristics

Patients with adenoid cystic carcinoma and non-adenoid cystic salivary gland cancer from the multicentric national DKTK MASTER program were included in the analysis. The DKTK MASTER program applies comprehensive molecular diagnostics to inform the care of adult patients with incurable cancers. DKTK MASTER inclusion criteria were advanced solid tumors of a rare histology or younger age (< 51y), no available standard therapy, and available fresh-frozen tumor tissue. The DKTK MASTER program was approved by ethics committees at all participating sites (Lead Heidelberg, S-206/2011). Written informed consent was obtained from all participants. Ethics approval for retrospective analysis of tumor samples from patients not participating in the DKTK MASTER program was obtained separately (Berlin, EA1/305/21). Clinical data was extracted from clinical documentation of the primary care facility detailing the patient history, as available.

### Immunohistochemistry

Formalin-fixed and paraffin-embedded (FFPE) surgical specimens of the patients were used to prepare slides for immunohistochemistry (IHC). IHC was performed on tissue microarrays (TMAs) with two cores for each case, according to standard procedures. As counterstaining Hematoxylin was used.

For the detection of the immune cells we used a polyclonal antibody against *CD3* for T-cells (solution 1:100, Dako) and monoclonal antibodies against *CD8* (clone *C8/144B*, solution, Dako) for T-helper cells, against *CD20* for B-cells (clone L26,solution 1:750, Dako), against CD68 for macrophages (clone PG-M1, solution 1:200, Dako), against *FOXP3* for T-regs (clone 236A/E7, solution 1:200, Abcam plc., Cambridge, UK) and *PD-L1* (clone E1L3N, solution 1:200, Cell signaling). All analyses were performed on Leica Bond Master.

For the detection of *VTCN1* we used the recombinant Anti-B7H4 antibody (clone EPR23665-20, solution 1:100, Abcam plc., Cambridge, UK). A positive expression for *CD3*, *CD20, CD68, CD8* was defined by a medium to strong intensity of membranous staining. A positive expression for *VTCN1* was defined by a medium to strong intensity of membranous staining of the tumor cells.

### Exome, Genome and bulk RNA seq sequencing

Bulk sequencing on fresh-frozen tumor tissue within the DKTK MASTER program was performed as previously described ^16^: DNA and RNA from tumor and DNA from matched blood samples were isolated using AllPrep Mini or Universal Kits (Qiagen). After library preparation (SureSelect Human All Exon, Agilent; TruSeq RNA Sample Preparation kit V2, Illumina), whole-exome and RNA-paired-end sequencing (2 × 151 bp; 2 × 101 bp) was performed with various HiSeq instruments (e.g HiSeq 4000 and NovaSeq 6000; Illumina).

### Exome, Genome and bulk RNAseq data processing

*Fastq* files were trimmed using *bbduk* by removing adapter sequences and then mapped using *bwa-mem* algorithm (version 0.7.17) onto the GRCh38 genome (GRCh38.d1.vd1, primary assembly with decoys and viral sequences). In case of RNA-seq data, alignment and gene expression quantification was performed with salmon (version 1.4.0, transcriptome version: GENCODE 33). Counts were normalized to gene length (TPM) and then used as gene expression measure. To compare gene expression between samples and to run GSVA, counts were “variance stabilized” via *vst* transformation using the *DeSeq2* package in R ^17^. Aligned exome and genome data was passed through a somatic variant calling pipeline. SNPs and short indels were detected by the *Mutect2* algorithm ^18^ using in-house panels of normals. The resulting vcf files were annotated via *jannovar* (version 0.26) and *vep* (version 102). Gene fusion products were detected with *arriba* ^19^ (version 2.3.0). Other dataset used here were retrieved from the TCGA database and the NCBI sequence read archive (SRA). For TCGA datasets *STAR* counts were used and transformed. Linxweiler et al ^13^ and Vos et al ^9^ comprised *fastq* files from SRA which were processed in the same manner as our data. Dou et al ^14^ provided only TPM values that were used directly.

### Bulk RNAseq downstream analysis

Differential expression was performed using the R package *DeSeq2* ^17^. For functional enrichment of DEGs reference gene sets available from MSigDB (release 2023 v1) and the package *tmod* ^20^ were used.

Sample-wise gene set enrichment was performed withGSVA implemented in the R package *gsva* ^21,22^. A set of several functional scores for immune infiltration published in four independent publications was used to cluster the patients. This set comprises a large 462 genes immune signature (IS) from Bindea et al, a large 186 genes T-cell signature (TIS) from Bindea et al, a 59 genes immune signature from Danaher et al, a 6 gene T-cell signature from Danaher et al, a 6 gene IFNG signature from Ayers et al, an 18 genes immune signature from Ayers et and a 7 gene APM (Antigen processing machinery) signature from Senbabaoglu et al 2016^23–26^. Furthermore, scores for several immune infiltrates were computed based on the set of 28 immune cell signatures from Senbabaoglu et al and based on the set of immune infiltrates signatures from Bindea et al ^23,24^.

As an additional validation other scores were computed: the cytotolic score ^27^, the ESTIMATE Immune score ^28^, the CIBERSORT absolute score ^29^. CIBERSORT was used both in absolute and relative mode to get the absolute and relative proportions of immune cells respectively. These analyses were performed using the *immunedeconv* package in R^30^ GSVA was applied using a gaussian kernel on variance stabilizing transformed counts (*vst*). The input for the *immunedeconv* methods (CIBERSORT, X-cell, ESTIMATE) were TPMs. Matrices with GSVA scores of immune-cells/infiltrates were clustered via hierarchical clustering with ward linkage and k=3. Immune infiltration scores were correlated against each other and the p-values were corrected for multiple testing. Association of IFNG score and several clinical parameters was computed with one-way and two-way anova models in R. To test differences between groups in any other settings a non-parametric, unpaired test such as Wilcoxon test was used. P-values were corrected using the holm-bonferroni method.

In the case of the integrated analysis with other cohorts, samples from tumor stage < 3 were filtered out (this filtering step was performed in order to analyze only advanced tumor samples) and the filtered combined expression matrix was batch corrected with the *limma* package in R before computing the GSVA scores.

Survival analysis was performed with R package *survminer* using only overall survival. Samples were divided into “high” or “low” for survival analysis by using the upper quartile and lower quartile of the respective immune score.

ACC1-ACC2 score was calculated based on *MYB-TP63* expression as described by Ferrarotto et al ^4^. If the score was > 0 the sample was labeled as ACC1, otherwise as ACC2.

Different entities were labeled as ID- or ED-derived based on a cut-off on median SOX10 expression and knowledge from literature, when applicable. ^31,32^

### WGS and WES downstream analysis

Annotated variants retrieved from exome and genome data were used for calculation of TMB, mutational landscape analyses and detection of mutational signatures. TMB was calculated as the number of non synonymous short variants per MB sequence. Variants used for TMB had at least 5% VAF, 10x coverage for the normal allele and 20x coverage for the mutant allele. VCF files were converted to maf files by the *vcf2maf* tool (version 1.6.21) and analyzed by the *maftools* package ^33^ in R. For mutational landscape analyses only mutations in coding sequence were used, whereas for mutational signatures all variants were used. The contribution to the 50 different SBS cosmic signatures for each sample was calculated using the extractSignatures and compareSignatures function. To test for positive selection the ratio of missense and synonymous mutations within the coding sequence of immunotherapy relevant genes was computed gene-wise. If the ratio > 1 there is a positive selection for the respective mutation(s). Tumor purity was assessed with ASCAT based on WGS and WES.^34^

### Tissue dissociation, nuclei preparation and single nuclei sequencing

Each sample of fresh frozen tumor tissue was suspended in NP-40 lysis buffer (10mM Tris-HCl (pH 7.4); 10mM NaCl; 3mM MgCl_2_; 0.01% NP-40; 1mM DTT; 2% BSA; 1U/µl RNAse inhibitor; Complete EDTA-free protease inhibitor) in a 1.5ml Eppendorf tube and disrupted with a plastic pestle. The suspension was incubated on ice for 5 minutes, filtered through a 70 µm pre-separation strainer and centrifuged at 4°C. The supernatant was removed and the nuclei were resuspended in nuclei wash buffer (PBS; 1% BSA; 0.4U/µl RNAse inhibitor). 3.5 µl DAPI were added, followed by incubation on ice. The mixture was filtered through a 40 µm Flowmi cell strainer and sorted with a 100 µm nozzle in an Eppendorf tube containing 200 µL sort buffer (PBS; 2% BSA; 2U/µl RNASe inhibitor). Single nuclei libraries were generated according to the Chromium Next GEM Single Cell 3ʹ Reagent Kits v3.1 (Dual Index) user guide (CG0003154) by 10x Genomics. Gel bead-in-emulsions (GEMs) were created using the Chromium Controller. In this step individual nuclei were encapsulated with a gel bead in a droplet, where barcoding occurs. The barcoded RNA was reverse transcribed into cDNA and amplified. The amplified cDNA underwent fragmentation, end-repair, A-tailing, adaptor ligation, and sample index PCR to create the final library. The prepared library was sequenced on a NovaSeq 6000 instrument (Illumina).

### Single nuclei data processing

Generated *fastq* files were aligned with *cellranger* (version v 7.1.0), which outputs count matrices for each sample. *cellbender* ^35^ was then used to filter count matrices and remove background noise. Expected cells and total droplets were estimated from cellranger quality control for each sample. The count matrices were further processed using the *scanpy* package in Python.

After cellbender filtering, additional filtering steps were performed. Only cells with at least 1000 UMIs and at least 300 expressed genes were kept. Cells with more than 20% mitochondrial and 10% ribosomal counts were removed. *scDBLFInder* was used for doublet detection ^36^, which was around 10% in the whole dataset.

Counts were normalized via size factor normalization ^37^. For PCA only 3000 to 5000 highly variable genes (depending on the analysis) were used. Furthermore, a total of 50 PCs were used for integration with *Harmony* ^38^. After assessment of 3 different integration methods, we found *Harmony* to be the best suited for our data. Harmony embeddings were used for knn-graph construction, UMAP and clustering. For clustering we used the *leiden* algorithm ^39^ with 0.5 resolution. Cell clusters were annotated manually based on markers retrieved from the literature ^40^ and the protein atlas ^41^ (proteinatlas.org). Immune cells were annotated automatically using published tools: *celltypist* ^42^ and *projecTILs* ^43^. Malignant cells were identified based on copy number variants (*inferCNV* of the Trinity CTAT Project. https://github.com/broadinstitute/inferCNV) and based on occurrence of somatic SNVs known from WES/WGS in the single-cell RNAseq reads ^44^. Malignant cells in ACC were further classified into myoepithelial- and luminal-like following a similar approach to the one used by Parikh et al ^45^. Luminal and myoepithelial scores were calculated and subtracted. If the difference was below or above a 0.3 threshold the cells were labeled either as luminal or as myoepithelial.

## RESULTS

### Cohort characteristics

A total of 104 patients with recurrent/metastatic salivary gland cancer from the DKTK MASTER program were analyzed. The most common tumor entity was adenoid cystic carcinoma (58%, 60/104 patients). In addition to ACC, further 12 tumor entities were included. 46% of patients were female and the median age was 52 years (IQR 41 - 61) at the time of tumor biopsy. One patient was younger than 18 years at time of diagnosis. The most common primary tumor site was the parotid gland in 47% of all samples. Two thirds of patients had received prior systemic therapy before inclusion in the DKTK MASTER program, with a median of one line (IQR 0 - 2) of therapy. Median duration of prior therapy lines was 139 days (IQR 57.5 - 153). A summary of clinical data is provided in **Figure 1** and **Table 1**.

**Figure 1:**
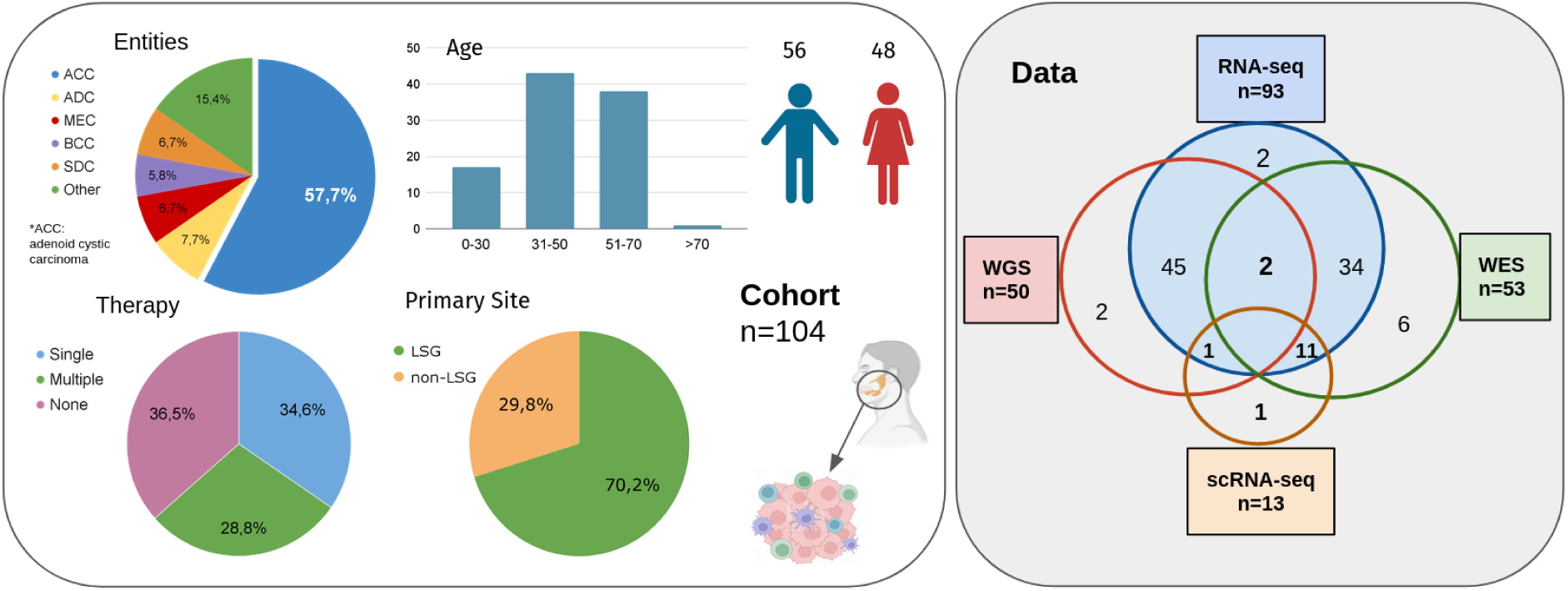
Study cohort and data. Different clinical characteristics of the patients are shown in the left panel. Right panel shows the different data layer and the overlaps in samples between them. Most samples (n=93) have bulk RNA seq. (*ACC= Adenoid Cystic Carcinoma, ADC: Adenocarcinoma, BCC=Basal Cell Carcinoma, MEC=Mucoepidermoid Carcinoma, SDC= Salivary Duct Carcinoma, LSG=Large Salivary Glands, WGS= Whole Genome Sequencing, WES=Whole Exome Sequencing*)

**Table 1:**
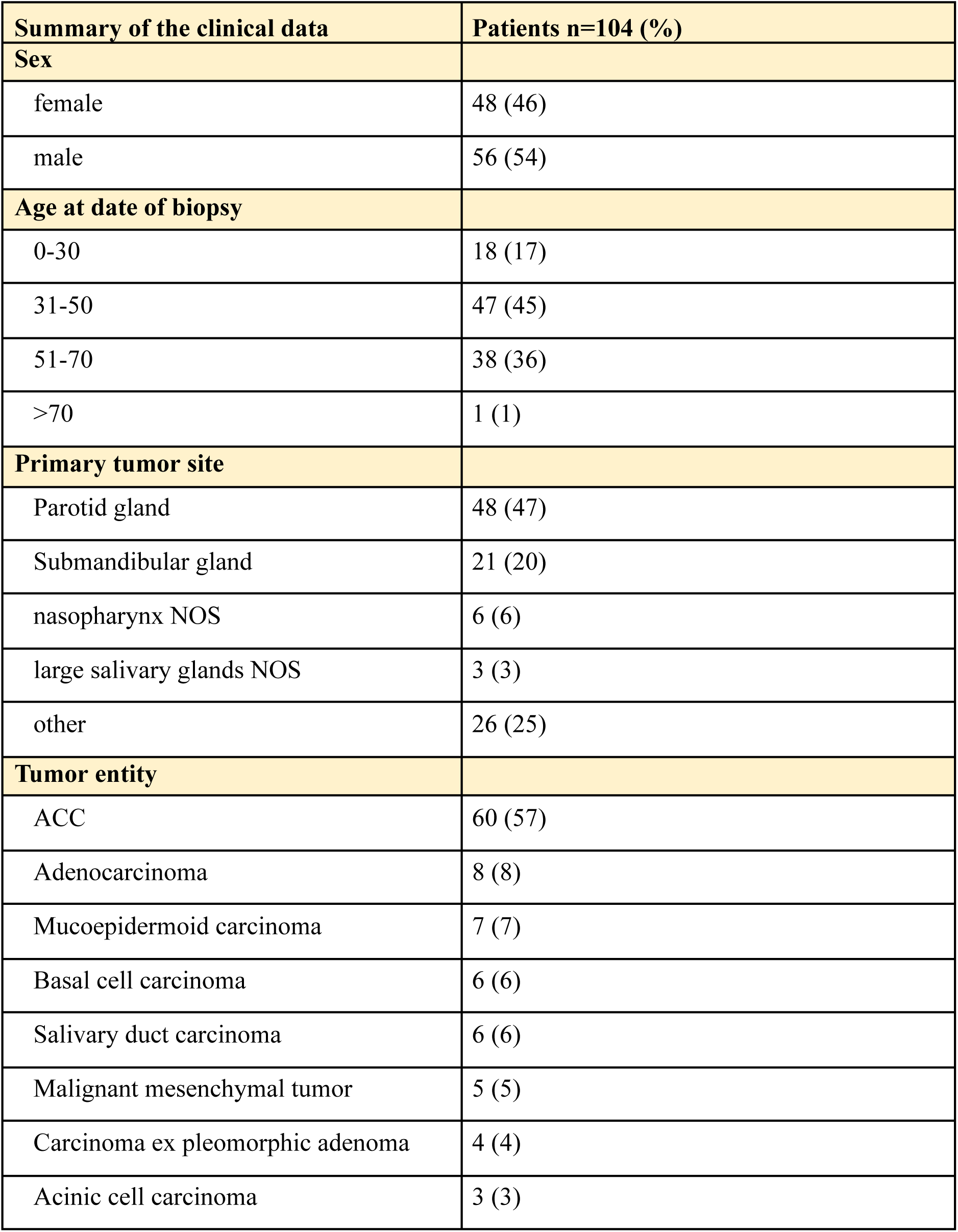
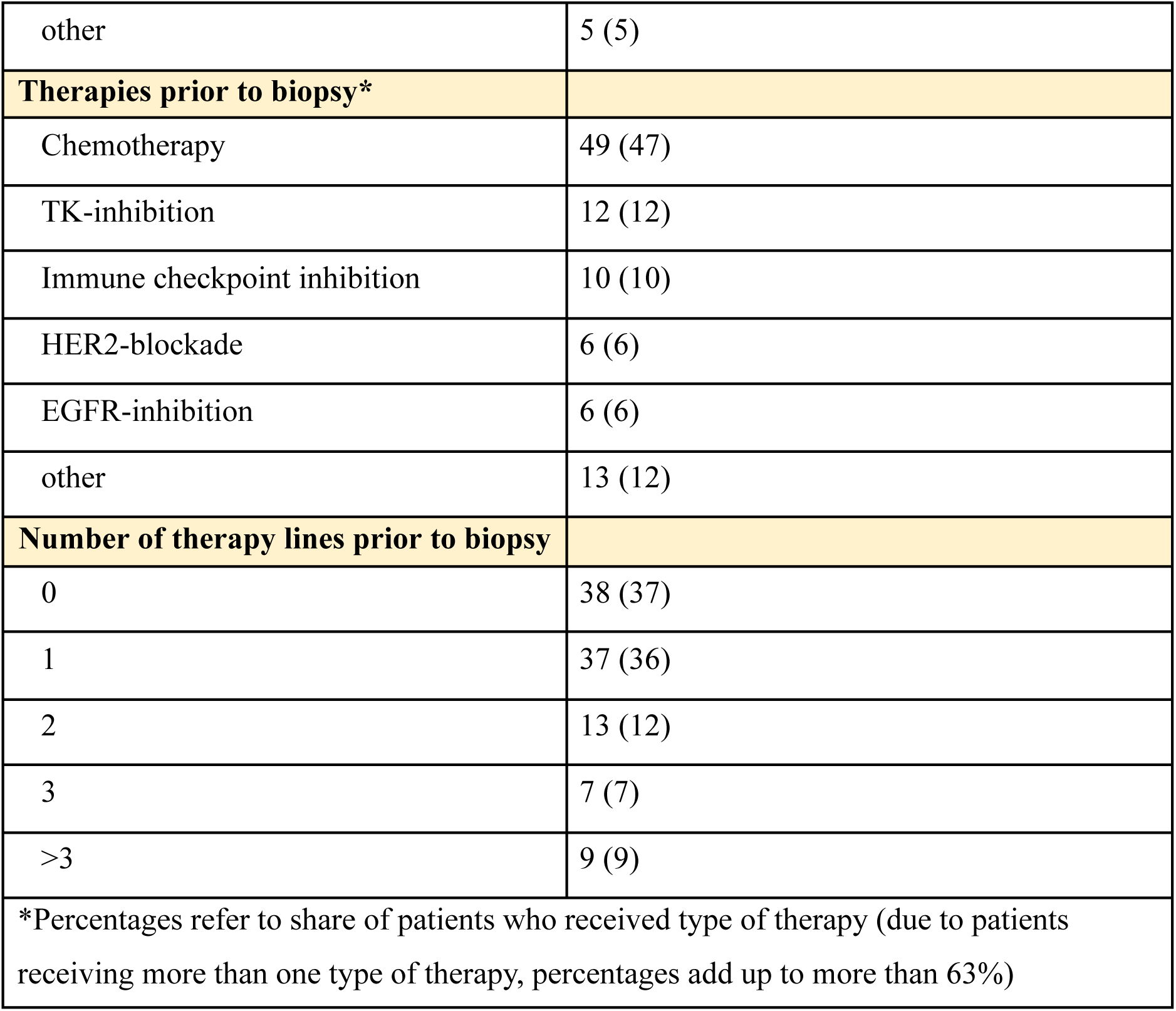
Baseline clinical characteristics of the cohort. *Due to rounding percentages may not add up to 100*.

| Summary of the clinical data | Patients n=104 (%) |
| --- | --- |
| **Sex** |  |
| female | 48 (46) |
| male | 56 (54) |
| **Age at date of biopsy** |  |
| 0-30 | 18 (17) |
| 31-50 | 47 (45) |
| 51-70 | 38 (36) |
| >70 | 1 (1) |
| **Primary tumor site** |  |
| Parotid gland | 48 (47) |
| Submandibular gland | 21 (20) |
| nasopharynx NOS | 6 (6) |
| large salivary glands NOS | 3 (3) |
| other | 26 (25) |
| **Tumor entity** |  |
| ACC | 60 (57) |
| Adenocarcinoma | 8 (8) |
| Mucoepidermoid carcinoma | 7 (7) |
| Basal cell carcinoma | 6 (6) |
| Salivary duct carcinoma | 6 (6) |
| Malignant mesenchymal tumor | 5 (5) |
| Carcinoma ex pleomorphic adenoma | 4 (4) |
| Acinic cell carcinoma | 3 (3) |
| other | 5 (5) |
| **Therapies prior to biopsy\*** |  |
| Chemotherapy | 49 (47) |
| TK-inhibition | 12 (12) |
| Immune checkpoint inhibition | 10 (10) |
| HER2-blockade | 6 (6) |
| EGFR-inhibition | 6 (6) |
| other | 13 (12) |
| **Number of therapy lines prior to biopsy** |  |
| 0 | 38 (37) |
| 1 | 37 (36) |
| 2 | 13 (12) |
| 3 | 7 (7) |
| >3 | 9 (9) |
| *Percentages refer to share of patients who received type of therapy (due to patients receiving more than one type of therapy, percentages add up to more than 63%) |  |

Bulk RNA-seq data was available for 93 patients (95 samples). Whole-Exome (WES) and Whole-Genome sequencing (WGS) data was available for 55 and 50 patients, respectively. Single-nuclei transcriptome sequencing was performed in selected samples (n=13), representing inflammation-high and inflammation-low in adenoid cystic and non-adenoid cystic histologies. A summary of molecular data layers is provided in Figure 1. Immunohistochemical validation analyses were performed in a tissue microarray of 13 samples.

We tested the samples for the presence of *MYB-NFIB* or *MYBL1-NFIB* fusion to confirm the pathological distinction of ACC. Around 65% of ACC had a *MYB-NFIB* and 4% a *MYBL1-NFIB* fusion, in concordance with the reported proportions from literature ^46^. None of the non-ACC samples had a *MYB-NFIB* fusion. Furthermore, principal component analysis in bulk RNA-seq data revealed most variance (14%) to be explained by tumor entity (*p=4.5e-10\*\*\**). No significant association could be found between other principal components and available clinical parameters. PC1, which separated ACC from other entities, was related to immune response (**Figure S1A**). Differentially expressed genes between ACC and non-ACC were also related to inflammation (*AUC=0.6, p.adj=2.4e-5\*\*\**), cell cycle (*AUC=0.8, p.adj=2.8e-10\*\*\**) and cell cycle in T-cells (*AUC=0.8, p.adj=1.5e-4\*\*\**) with lower expression in ACC (**Figure S1B**).

### Advanced salivary gland cancers cluster into 3 groups of immune infiltration

To further analyze tumor inflammation in advanced SGC, bulk gene expression data from 95 samples (93 patients) was analyzed. Seven different published functional inflammation signatures were used to identify samples with an inflamed TIM (**Figure 2B**). A primary analysis of TIM composition was performed using scores for 2 different sets of signatures of immune infiltrates. Clustering on these scores was in agreement with the clustering based on functional scores. Other measures of immune infiltration besides the 7 signatures shown in Figure 2B were computed. All of the mentioned immune scores showed significant correlation between each other after multiple testing corrections (**Figure 2A**).

**Figure 2:**
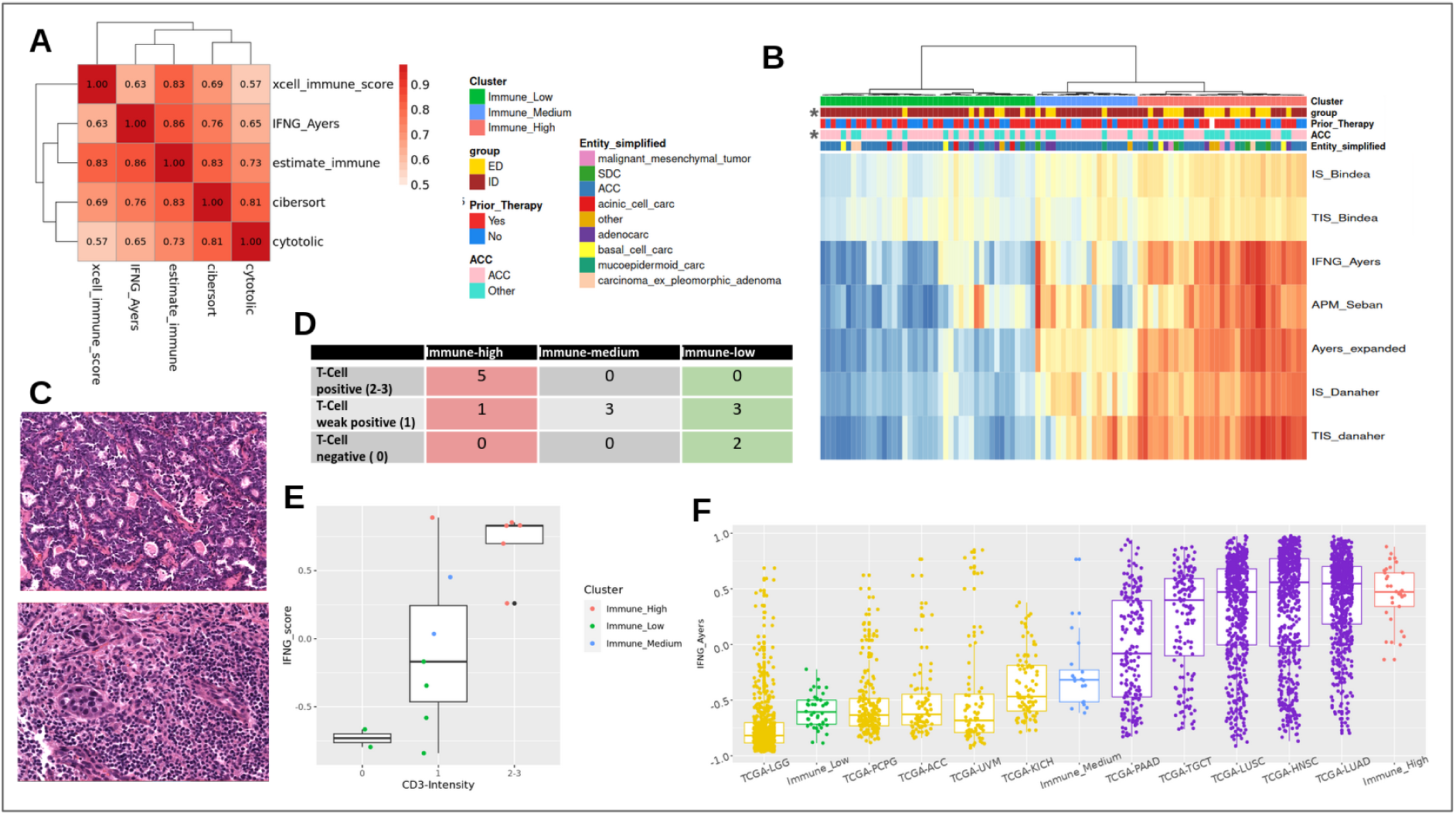
Validation of immune clusters. **A:** Correlation heatmap between different scores and methods (all correlations are significant after p value correction) **B:** Heatmap of GSVA scores of 7 immune signatures (see methods) for all samples (n=95). Samples are clustered by hierarchical clustering and annotated by entity, group (ID/ED) and therapy status. **C:** Example of T-cell negative (above) and T-cell positive (intensity=3) sample.D: Results of IHC for pan T-cell marker (*CD3*) The numbers in the table represent the amount of samples belonging to each category. **E:** Intensity of pan T-cell marker versus IFNG bulk score. **F:** Comparison of immune clusters to TCGA most (PAAD, TCGT, LUSC, HNSC, LUAD) and least inflamed cohorts (LGG, PCGP, ACC, UVM, KICH)

Hierarchical clustering based on GSVA scores identified 3 clusters of immune-infiltration (**Figure 2B)** with around 34% and 44% of samples belonging to the immune-high and immune-low clusters, respectively. ACC were significantly under-represented in the immune-high cluster in GSVA based results (*Fisher test, p=0.002**, OR=0.3*). Yet, we identified a small subset of ACC samples (n=10, ∼17%) with high immune infiltration, which could also be validated in a separate analysis in only ACC samples (n=57).

An integrated analysis with previously published SGC datasets ^9,13,14^ further confirmed 3 clusters of immune infiltration with ACC being significantly underrepresented in the immune-high group. Importantly, a small immune-high ACC subgroup could be validated in all 3 cohorts (**Figure S2B**). Compared to pan-cancer TCGA data, inflammation (IFNG score) in the ACC cohort was comparable to those of the 5 least inflamed TCGA cohorts. Other entities showed a much higher variance in their inflammation score, however inflammation scores of immune-high samples aligned with the 5 highest inflamed TCGA cohorts (**Figure 2F**).

Immune clusters were validated immuno-histochemically in a tissue microarray of 14 samples with available bulk sequencing data (**Figure 2C-E**). CD3 staining intensity was significantly correlated with the IFNG score (*rho=0.74, p=0.002\*\**) (**Figure 2E**).

### Correlates of tumor inflammation in advanced SGC

We tested for association of different clinical parameters with the immune clusters and inflammation scores. Apart from ACC-histology (p.adj= 0.002 **) and ID-group (p.adj= 0.00012 ***) no significant associations were found for sample origin (metastatic/primary), primary site (large salivary glands/other), age, cohort batch, sex, and therapy status for several therapies (received therapy/therapy-naive) after p-value correction. The association with tumor entity (ACC vs non-ACC) remained significant after correcting for almost all other confounders, except for the biopsy site. In ACC, inflammation was higher in samples from metastatic origin compared to samples from primary tumors, which was not observed in other entities (**Figure S3A-B**). Nevertheless, inflammation was lower in ACC when looking at all sites of biopsy, confirming the entity as being the most important factor influencing inflammation. Among all sites of biopsy only primary and lung samples were evaluable for inflammation, other tissues could not be tested due to the low number of samples.

TMB was calculated based on WES and WGS data and was significantly lower (*Wilcoxon test, p=0.007\*\** and *p=0.0004\*\*\**) in ACC than non-ACC samples (**Figure S4A**). The TMB of ACC was also in the lower spectrum when compared to other TCGA entities (8/33, **Figure S4B**). TMB and IFNG score showed a weak positive correlation (*rho=0.31, p=0.002\*\**, **Figure S4C**) in the combined WES and WGS data (n=103). This correlation was mostly driven by non-ACC samples (Other, **Figure S4D**). Multivariate analysis showed no relevant influence of other clinical covariates on this association. Tumor purity was computed based on WGS/WES data (ASCAT). Inflammation was significantly negatively correlated with tumor purity, but with a rather low correlation coefficient (0.4).

No positive selection could be identified in mutations affecting immune checkpoints and other immunotherapy-relevant genes ^47^. Mutational signatures ^48^ were extracted for WGS data. Four main signatures could be found in 52 patients with WGS including APOBEC-signatures (SBS2,13), chemotherapy-related signature (SBS31), clock-like signature (SBS5) and signature of unknown etiology (SBS17). APOBEC signature was prevalent in only 2 samples (one of them representing a rare case of HPV-positive ACC). Therefore, no association could be found between APOBEC mutagenesis and inflammation subgroups. Around 77% of samples had a prevalent SBS5-like signature, which is defined as being likely related to age or to DNA repair deficiency in cancer. None of the signatures were associated with inflammation in the samples (data not shown).

Since TMB was not associated with inflammation in ACC samples, we tested for the association of MYB-fusion, NOTCH mutations and ACC subtype score ^4^ with inflammation in ACC samples separately. The ACC score was negatively correlated with immune scores (*rho ∼ −0.3, p ∼0.03\**). A significant association with the antigen processing machinery (APM) was identified in these samples (*rho=-0.49, p=0.0002\*\*\**, **Figure S3C**). The negative correlation indicates that samples belonging to ACC1 subgroup tend to have a lower immune infiltration, which might be mediated by deficient antigen processing. ACC1 samples were found to have a significantly worse prognosis than ACC2 (**Figure S3D**).

### TIM composition analysis reveals Macrophage-dominance in advanced SGC

To identify the TIM cell composition in advanced SGC, single nuclei RNA-seq data was analyzed for 13 samples. The single nuclei cohort comprised 5 ACC samples (1 primary site, 4 metastatic sites) and 6 other entities (2 basal cell carcinoma, 1 carcinoma ex pleomorphic adenoma, 2 adenocarcinoma NOS, 1 carcinosarcoma, 1 mucoepidermoid carcinoma, 1 salivary duct carcinoma). After quality control and filtering the total amount of cells was 85142.

**Table 2** shows the sample origin, tumor entity and several quality metrics for each sample sequenced. Data analysis revealed 23 different cell clusters, which could be assigned to 6 major cell types: fibroblasts, endothelial cells, immune cells, alveolar cells (present only in the lung metastases) and malignant cells (**Figure 3A-B** and **Figure S5A-C**). More than half of all identified immune-cells came from 2 lymph node metastases (**Figure 3A**, P-2 and P-8). Therefore, these samples were removed to further analyze TIM composition. The majority of immune cells were labeled as macrophages (mean: 60% sd: 25), followed by T-cells (mean: 22%, sd: 18) and dendritic cells (median: 6%, sd: 5) (**Figure 3C-D**). The least abundant immune cells were B-cells, which were almost only present in lymph node metastases. No association between overall abundance of immune cells and T-cell proportions could be identified in the single nuclei cohort. Samples labeled as immune high from bulk data analysis had more immune cells than those labeled as immune low or medium (with the exception of one sample). Immune high samples had a median percentage of ∼14% immune cells (2 lymph node metastasis with >48% immune cells), whereas immune low and medium together had a median of ∼4% of immune cells. ACC also had an overall lower immune infiltration with a median of ∼4% compared to non-ACC (without lymph node metastases) with a median of ∼7%. Immunohistochemical analysis of macrophage distribution in SGC (n=3) revealed clustering of CD68-positive cells at the tumor margin and co-localization with T-cells (**Figure 4E)**

**Figure 3:**
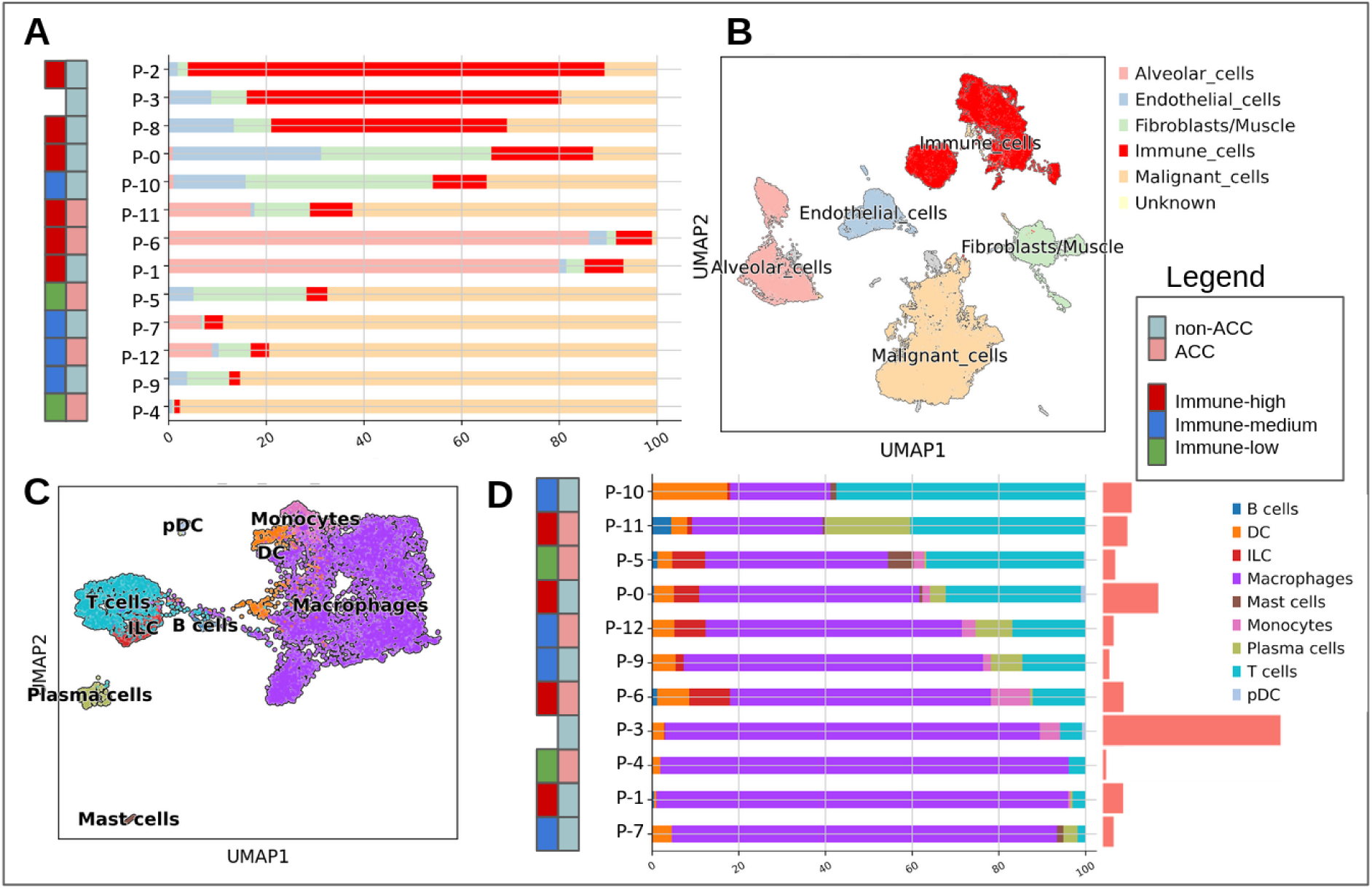
TIM composition analysis based on single nuclei data. **A:** Proportions of major cell-types in single nuclei data (for cluster names see panel B). Samples are annotated by tumor entity and bulk-based immune cluster (see legend). P-3 does not have available bulk data and therefore lacks immune cluster annotation. **B:** UMAP plot of integrated data, annotated by major cell types. **C:** UMAP of immune cells (n=11) annotated by celltypist [cit]. **D:** Proportions of immune cell types in single nuclei data (for cluster names see panel B). Samples are annotated by tumor entity and bulk-based immune cluster (see legend). The bars on the right represent the proportion of immune cells in the sample.

**Figure 4:**
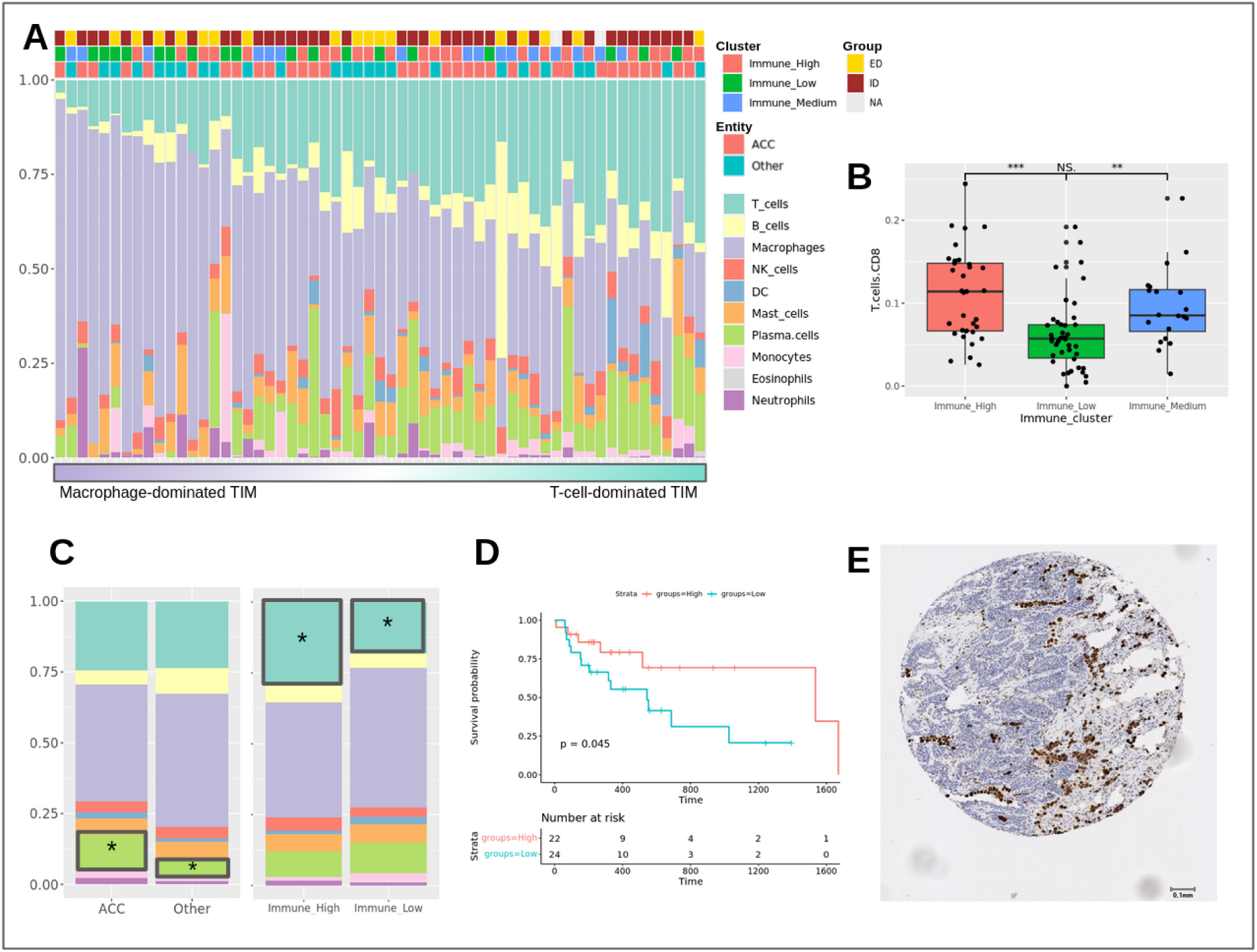
TIM composition analysis based on subset of bulk RNA-seq data (n=59). **A:** Deconvoluted proportions of immune cell types in the cohort (n=59). Samples are sorted by macrophage to T-cell ratio. Samples are annotated by immune cluster,entity and group. **B:** Predicted relative proportions of CD8 T-cells in 3 immune clusters. and **C:** Proportions of immune cells grouped by entity (left) and immune cluster (right). **D:** Survival plots (Overall Survival) of samples with high and low T-cell proportion. **E:** Representative CD68 staining shows clusters of macrophages.

**Table 2:** Single nuclei samples characteristics and quality metrics. *(ACC=Adenoid Cystic Carcinoma, AdC=Adenocarcinoma, BCC=Basal Cell Carcinoma, CS=Carcinosarcoma, CEPA=Carcinoma ex Pleomorphic Adenoma, MEC=Mucoepidermoid Carcinoma, SDC=Salivary Duct Carcinoma)*

| Sample | Site Biopsy | Entity | # nuclei | median UMI counts / nucleus | median transcripts / nucleus |
| --- | --- | --- | --- | --- | --- |
| P-1 | lung | CS | 6640 | 4254 | 2235 |
| P-2 | lung | AdC | 2774 | 1740 | 1108 |
| P-3 | lymph node | SDC | 4344 | 5624 | 2277 |
| P-4_1 | primary | CEPA | 3957 | 5034 | 2427 |
| P-4_2 | primary | CEPA | 3839 | 4824 | 2395 |
| **P-5** | primary | **ACC** | 4667 | 2495 | 1545 |
| **P-6** | skin | **ACC** | 5455 | 3867 | 2064 |
| **P-7** | lung | **ACC** | 10273 | 3750 | 1892 |
| P-8_1 | lung | BCC | 8082 | 7149 | 2834 |
| P-8_2 | lung | BCC | 7864 | 7152 | 2773 |
| P-9_1 | lymph node | BCC | 5973 | 4756 | 2292 |
| P-9_2 | lymph node | BCC | 4664 | 3961 | 2049 |
| P-10 | primary | MEC | 4802 | 2179 | 1358 |
| P-11 | skin | AdC | 2861 | 3261 | 1811 |
| **P-12** | lung | **ACC** | 6497 | 2920 | 1804 |
| **P-13** | lung | **ACC** | 4343 | 4095 | 2277 |

Bulk based scores, in particular IFNG score were in agreement with immune cell proportions from single nuclei data (*rho=0.8, p=0.002\*\**). Among several deconvolution methods, CIBERSORT showed the best agreement with single cell based results and IHC. We therefore performed a deconvolution analysis (CIBERSORT) on bulk data to explore the TIM in a larger number of samples. 59 samples had significant results from CIBERSORT and could be used for subsequent analyses. Similar to single nuclei based results, most of the analyzed samples had a myeloid-dominant microenvironment (**Figure 4A**). On average, myeloid cells made up 55% (41% Macrophages) and T-cells 25% of the TIM in the entire cohort. The relative proportion of CD8 T-cells was slightly higher in the immune-high group, compared to the other two groups (*Wilcoxon test*, *p=1.7e-4\*\*\**)(**Figure 4B)**. The relative proportions of different immune cell types, except plasma cells, did not differ significantly between ACC and non-ACC after multiple testing corrections (**Figure 4C)**. The proportions of the 8 different immune cell types did not cluster by immune cluster assignment nor by tumor entity (**Figure 4A-C)**. When comparing the T-cell to Macrophage ratio of this cohort to several TCGA and another advanced SGC cohort ^49^, we could confirm the TIM of advanced SGC to be Macrophage-dominated. The T-cell to Macrophage ratio in SGC was among the 3 lowest, comparable to the one of TCGA-LGG. The median T/Macrophage ratio of both analyzed SGC cohorts was below 1.

Survival analysis showed an association between a high T-cell relative proportion **(Figure 4D)** as well as a high plasma cell relative proportion and better overall survival. After p-value correction however, only the association with plasma cells remained significant. On the other hand, no significant effect of macrophage proportion was found on overall survival, although a tendency to shorter survival in samples with high macrophage content was observed (data not shown).

Abundance of macrophages and T-cells were correlated inversely both in bulk data (*rho=-0.57,p=5e-09\*\*\**) as well as in single nuclei data (*rho=-0.91,p=1e-05\*\*\**). Also, the majority of macrophages were predicted as being M2 polarized (median 81% of all macrophages). Both points indicate an immune-suppressive role of macrophages in the TIM.

### Analysis of biomarkers for immunotherapy

Among 22 evaluable patients treated with immune checkpoint inhibitors, only one patient (adenocarcinoma NOS) achieved clinical benefit (time on treatment 1679 days). CIBERSORT analysis of TIM composition revealed a high T-cell to macrophage ratio in this patient compared to other evaluable samples (**Figure 5A**, marked with a pink rectangle). Independent validation in a second cohort ^5^ of patients with pretreatment sequencing before immune checkpoint inhibition also revealed clinical benefit in the patient with the highest T-cell infiltration (**Figure 5B)**. Both patients had a moderate immune-cell infiltration in the context of the whole cohort, indicating that the T-cell relative proportion rather than the overall immune infiltration might be of importance for the ICI-response.

**Figure 5:**
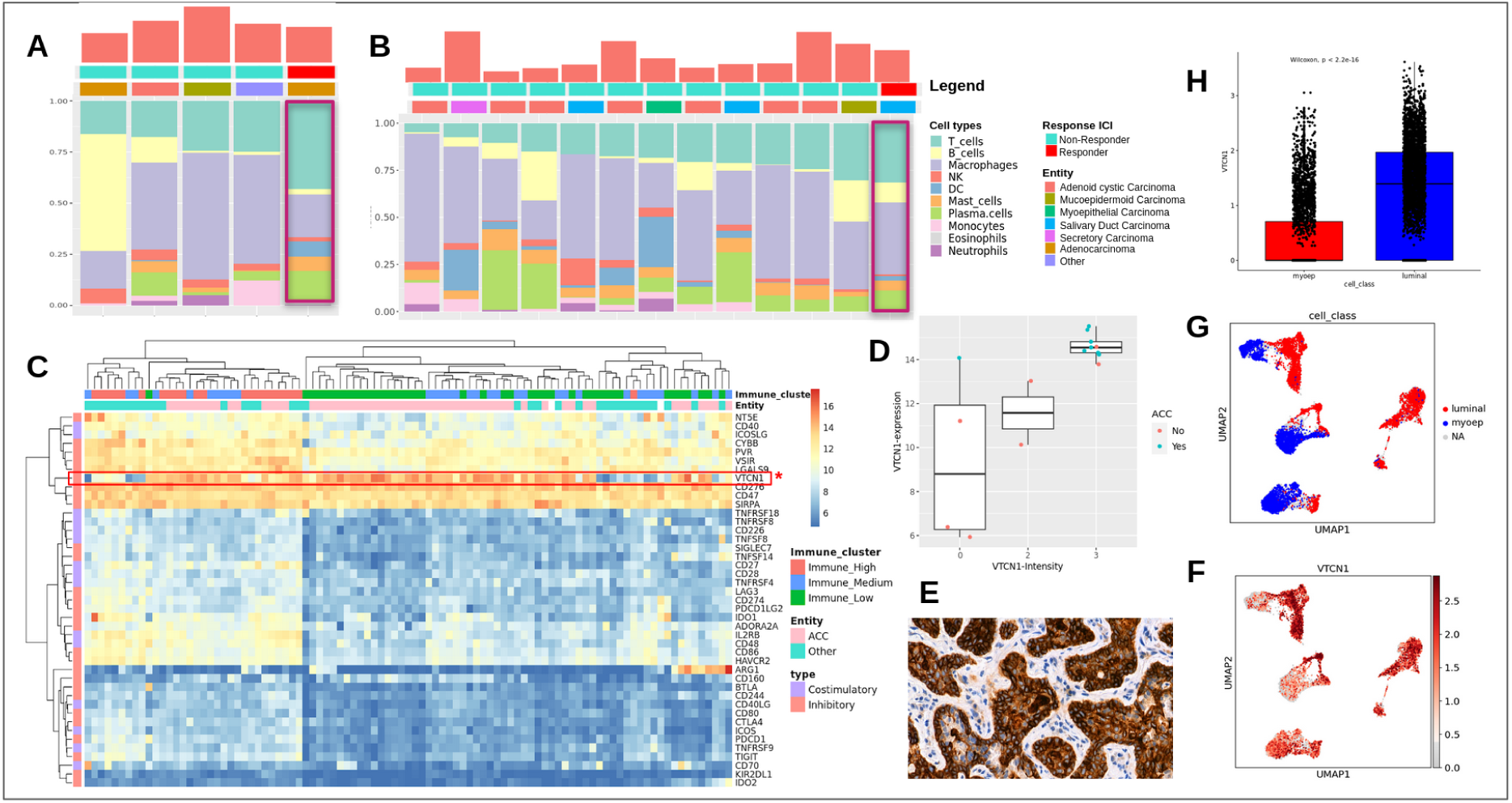
Analysis of biomarkers for immunotherapy. **A:** Deconvolution results (p<0.05) of 5 samples who received ICI. **B:** Validation of results from Figure E in an independent cohort (Vos et al) of SGC samples treated with ICI. Responders were defined as SD >6 months or PR/CR. **C:** Expression matrix based on variance-stabilizing transformed counts of selected immune checkpoints. The checkpoints are annotated on the left as co-inhibitory or co-stimulatory. The samples are annotated based on the tumor entity and the immune - cluster assignment. *VTCN1* is highlighted in red. **D:** VTCN1 RNA expression grouped by IHC staining intensity **E:**. Exemplary staining of *VTCN1* in an ACC sample reveals strong staining intensity on tumor cells. **F:** Expression of *VTCN1* in ACC malignant cells. UMAP shows non-integrated data. Cells cluster by donor. **G:** UMAP of malignant cells in ACC, same as in G but colored by assigned cell type (myoepithelial- or luminal-like). **H:** Boxplot with expression of *VTCN1* in myoepithelial- vs luminal-like malignant cells showing significantly higher expression in luminal ACC cells.

To identify additional treatment targets, a set of 43 immune checkpoint molecules was analyzed for differential expression **(Figure 5C)**. Most of the immune checkpoints were enriched in the immune-high cluster. We could observe an overexpression of inhibitory members of the immunoglobulin superfamily *VTCN1* (syn: B7-H4, *L2FC= 2.2, p.adj=2e-08\*\*\**) and *CD160* (*L2FC= 3.5, p.adj= 2e-19\*\*\**) in ACC compared to other entities. Several immune checkpoints were significantly under-expressed in ACC (*p.adj < 0.001*** and L2FC < −1.5*), such as inhibitory members of the immunoglobulin superfamily *PD-1,* its ligand *CD274,* and *TIGIT,* the co-stimulatory molecules *TNFSF8* and *TNFRSF9 (4-1BB),* and *NT5E (CD73),* which converts extracellular ATP to immunosuppressive adenosine and seems to modulate specifically the TAM polarization.

We additionally analyzed mRNA expression of a comprehensive list of potential targets of T-cell-receptor based immunotherapy strategies. Expression of any potential treatment target was identified in the majority of samples. Expression clusters of target groups were identified, e.g. *CTAG1B* (*NY-ESO-1*) and *MMP7* were found to be significantly overexpressed in ACC (*L2FC=4.7, p.adj=1.9e-10\*\*\** and *L2FC=3, p.adj=1.2e-11\*\*\**). Around 25% of the cohort had high expression of some or multiple genes of the MAGE family (**Figure S6**).

Protein expression of *VTCN1* was validated immunohistochemically in 14 samples and correlated with bulk RNA expression (**Figure 5D,E**). *VCTN1* expression was further confirmed in tumor cells of ACC samples in single nuclei RNA seq data. No association between *VTCN1* expression and different immune clusters in ACC was seen. Also, *VTCN1* expression was only weakly correlated with ACC subtype and worse survival in ACC samples (n.s.). Although ID-derived histologies had a higher *VTCN1* expression when compared to ED-histologies, *VTCN1* was still significantly over-expressed in ACC samples when compared to other ID-derived histologies (Wilcoxon, p=1.5e-3**). This indicates that *VTCN1* is a suitable marker for ACC specifically. Among other ID-derived histologies, basal cell carcinoma and carcinoma ex-pleomorphic adenoma were those with the highest VTCN1 expression. Further analyses in single nuclei data demonstrates that VTCN1 is over-expressed in luminal-like cells and less expressed in myoepithelial-like cells within ACC samples (**Figure 5F-H**). ACC1 (n=1) had almost exclusively luminal-like cells, whereas ACC2 (n=3) had more myoepithelial-like cells. This might be the reason why ACC1 samples have a higher *VTCN1* expression than ACC2 samples^50^.

## DISCUSSION

Effective immune therapies are lacking for patients with advanced salivary gland cancer. Response rates with PD-1 or PD-1/CTLA-4 directed therapies are low and range from 4-16% in prospective clinical trials ^9,51^. Consequently, the use of immune checkpoint inhibitors has not been recommended outside of clinical trials in current guidelines ^12^. In order to identify potential biomarkers and therapeutic strategies for immunotherapy in these hard-to-treat tumors, we analyzed a large cohort of advanced salivary gland cancer using bulk and single-cell sequencing data.

An analysis of advanced tumors is relevant, since they represent a potential intention-to-treat cohort and might differ biologically from early-stage tumors, which are usually overrepresented in genomic tumor analyses. Using this cohort, we were able to show that no relevant differences of inflammation status exist between early and late stage disease. However, in ACC a higher inflammation was observed in lung metastases when compared to primary tumors. These results have to be interpreted with caution with regards to the small sample size. However, pulmonary metastases have been associated with a more favorable clinical course in ACC ^52^ and are likely linked to different ACC subgroups.

Results show significantly less tumor inflammation in ACC compared to non-adenoid cystic carcinoma histologies, in line with previous analyses in early SGC ^13,14^. These results highlight the heterogeneity between tumor subtypes in SGC, which was also shown in a reanalysis of early-stage SGC gene expression data, showing an association between cell-of-origin and tumor inflammation^32^. Several mechanisms might contribute to these phenotypic differences. Inflammation was associated with tumor mutational burden mostly in non-ACC rather than ACC histologies in our analyses, albeit the effects were marginal which might be related to an overall lower tumor mutational burden in the majority of samples. In ACC, inflammation was associated with ACC subtypes, which have previously been suggested to be associated with prognostic relevance ^4^. Our results suggest that differences in the antigen processing machinery (APM) might contribute to these differences in inflammation between ACC 1 and ACC 2.

Identification of immune cell compositions from bulk data is challenging. Using single-cell transcriptome validated bulk deconvolution analyses, we were able to identify a myeloid compartment defining both ACC- and non-ACC salivary gland cancer TIM. Macrophages have been characterized as drivers of immune-suppression and resistance to immune checkpoint inhibition ^53^. The high macrophage content, in particular of M2-polarized macrophages in advanced salivary gland cancers, might therefore mediate immune checkpoint inhibitor resistance in these patients despite the presence of an inflamed TIM. The comparative analyses with TCGA data show that a relative macrophage predominance is especially relevant in SGC.

Interestingly, a high T-cell to macrophage ratio was associated with response to immune checkpoint inhibitors in individual patients benefiting from immune-checkpoint inhibition in our and a second validation cohort ^9^. In both cohorts only a subset of samples could be confidently analyzed via deconvolution, thus limiting statistical power. Yet, these results merit further validation of T-cell to macrophage ratio as a potential predictive biomarker for immune checkpoint inhibition in advanced SGC. Also, further analyses to elucidate the role of macrophages in ICI response are needed, as they are the most abundant cell-type in the TIM and might modulate T-cell infiltration. Macrophage-directed agents should be further investigated either alone or in combination with other immunotherapies for patients with SGC.

High expression of *VTCN1* (B7H4) was identified in ACC and might represent an additional treatment target, thus validating previous findings in cohorts with mostly earlier-stage ACC ^15^. In contrast to these previous results, we only found a minor difference in *VTCN1* expression between ACC-1 and -2 subtypes, which might therefore be less pronounced in more advanced disease stages. However, significantly different *VTCN1* expression was identified between luminal- and myoepithelial cells within adenoid cystic carcinoma, which might impact therapeutic efficacy of *VTCN1*-directed agents and underlie previously reported differences between ACC subtypes.. *VTCN1* has been shown to negatively regulate T-cell immune response ^54^ and was negatively associated with tumor-infiltrating lymphocytes and *PD-L1* expression in breast cancer ^55^. Hence, over-expression of *VTCN1* on ACC cells could explain the rather low immunogenicity in ACC and poor response to ICI and might represent a novel treatment target for advanced ACC, irrespective of ACC subtype. In addition to immune checkpoints, expression of target antigens for novel immunotherapy strategies was identified in a relevant subset of samples, including a histology-predominant expression of *NY-ESO1* in ACC, in line with previous reports ^56^. Advanced SGC should therefore be incorporated in ongoing trials, such as T-cell-receptor based immunotherapies.

In conclusion, an inflamed TIM can be observed in a subset of advanced SGC with ACC showing significantly less inflammation. TMB in non-ACC and antigen processing in ACC might contribute to these observed phenotypes. Novel immune checkpoint inhibitors and T-cell-receptor based therapies should be further investigated in biomarker-stratified SGC. Among these targets, significant *VTCN1* overexpression in advanced ACC might represent a novel treatment option. A clinical trial of a *VTCN1*-directed therapy is ongoing (NCT05194072). Furthermore, TIM cell compositions are characterized by macrophage predominance, representing an additional potential treatment target, whereas a high T-cell/macrophage ratio should be further investigated as a predictive biomarker for immune checkpoint inhibitors. These results further support the development of biomarker-based immunotherapy strategies in advanced SGC.

## List of Abbreviations

ACC: adenoid cystic carcinoma
ADC: adenocarcinoma
APM: antigen processing machinery
AUC: area under the curve
BCC: basal cell carcinoma
CEPA: carcinoma ex pleomorphic adenoma
CS: carcinosarcoma
DEG: differentially expressed genes
FFPE: formalin-fixed paraffin-embedded
GSVA: gene set variation analysis
HPV: human papilloma virus
ICI: immune checkpoint inhibition
IHC: immunohistochemistry
IS: immune signature
LSG: large salivary gland
MEC: mucoepidermoid carcinoma
NOS: not otherwise specified
ORR: overall response rate
PCA: principal component analysis
PR: partial response
SDC: salivary duct carcinoma
SBS: single base substitution
sd: standard deviation
SGC: salivary gland cancer
ssGSEA: single sample gene set enrichment analysis
TIM: tumor immune microenvironment
TMB: tumor mutational burden
WES: whole-exome sequencing
WGS: whole-genome sequencing
PAAD: pancreatic ductal adenocarcinoma
TCGT: testicular germ cell tumors
LUSC: lung squamous cell carcinoma
HNSC: head and neck squamous cell carcinoma
LUAD: lung adenocarcinoma
LGG: low grade glioma
PCGP: Pheochromocytoma and Paraganglioma
UVM: uveal melanoma
KICH: kidney renal clear cell carcinoma

## Declarations

### Ethics approval and consent to participate

The DKTK MASTER program was approved by ethics committees at all participating sites (Lead Ethics Committee Heidelberg, S-206/2011). Written informed consent was obtained from all participants. Ethics approval for retrospective analysis of tumor samples from patients not participating in the DKTK MASTER program was obtained separately (Ethics Committee Charité Berlin, EA1/305/21).

### Consent for publication

Not applicable

### Availability of data and materials

The datasets generated and/or analysed during the current study are not publicly available due to missing approval by participating patients but are available from the corresponding author on reasonable request.

### Competing interests

CH received honoraria, research funding and/or consulting/advisory board from Roche, Novartis, Boehringer Ingelheim. SF has had a consulting or advisory role and received honoraria, research funding, and/or travel/accommodation expenses funding from the following for-profit companies: Amgen, AstraZeneca, Bayer, Eli Lilly, Pfizer, PharmaMar, and Roche. DTR has received honoraria, research support and/or travel/accommodation expenses from Bayer, Eli Lilly, Bristol-Myers Squibb, Roche and Seagen.

### Funding and Support

The MASTER program is supported by the NCT Molecular Precision Oncology Program, DKFZ, and DKTK. This project was supported by the Deutsche Forschungsgemeinschaft (No. RTG2424 CompCancer, EZ) and a Berlin Institute of Health Booster Grant (EZ, DB, DTR). DTR is a participant in the Berlin Institute of Health - Charité Clinical Scientist Program funded by the Charité - Universitätsmedizin Berlin and the Berlin Institute of Health. Part of this work was supported by the ACC Research Foundation (DTR).

### Author’s contributions

EZ conceptualized and performed analyses, interpreted the data and drafted the manuscript; BvdE performed analyses and data acquisition and drafted the manuscript; IP performed analyses and interpreted data and drafted and revised the manuscript; ACAP interpreted the data and revised the manuscript; KK acquired and interpreted data and revised the manuscript; AM performed analyses, interpreted data and revised the manuscript; PH acquired and interpreted data and revised the manuscript; CH, FK, IP, MB, CHB, SK, MVT and DH acquired data and revised the manuscript; LGTM interpreted data and revised the manuscript; UK interpreted data and revised the manuscript; HG and SF acquired data and funding, designed the work and revised the manuscript; SO acquired data and revised the manuscript; UK designed the work, interpreted data and revised the manuscript; EB, DB and DTR contributed to the conception and design of the work, interpretation and acquisition of data, acquisition of funding and drafted and revised the manuscript.

## Acknowledgments

The authors thank the NCT/DKFZ Sample Processing Laboratory, the DKFZ Next-Generation Sequencing Core Facility, and the DKFZ Omics IT and Data Management Core Facility for technical support as well as the Core Unit Genomics - Single-Cell Technologies, Max Delbrück Center, Berlin. The results published here are in part based upon data generated by the TCGA Research Network: https://www.cancer.gov/tcga.

## Supplementary figures

**Figure S1.**
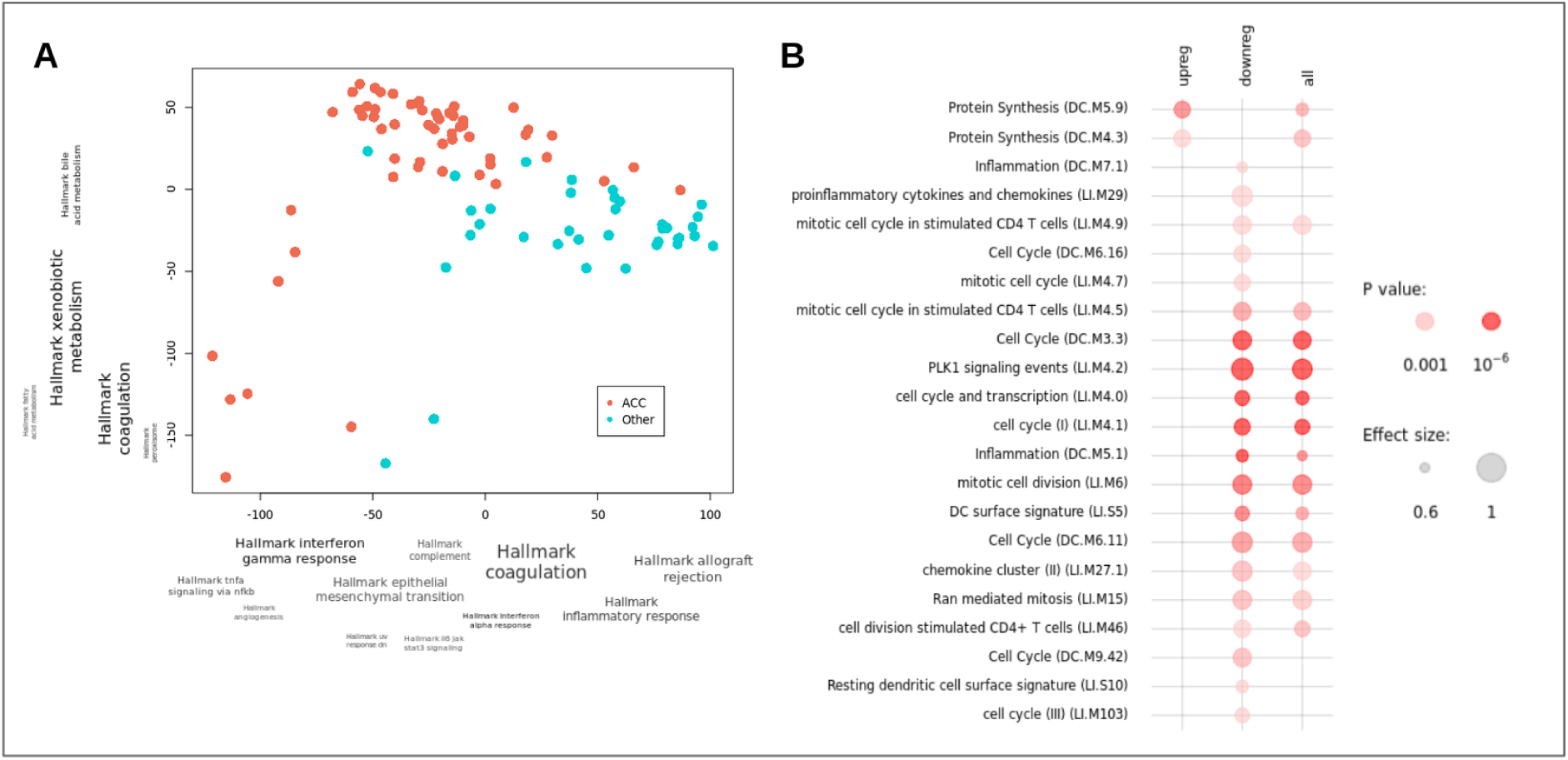
**A:** PCA plot of transcriptome. Axes are annotated by functional enrichment (HALLMARK) of genes contributing to PC1 and PC2. Samples are colored by tumor entity (ACC vs Other). **B:** Functional enrichment of differentially expressed genes between ACC and other SGC. First column represents genes upregulated in ACC, the mid column genes that are downregulated in ACC and right all DEGs

**Figure S2.**
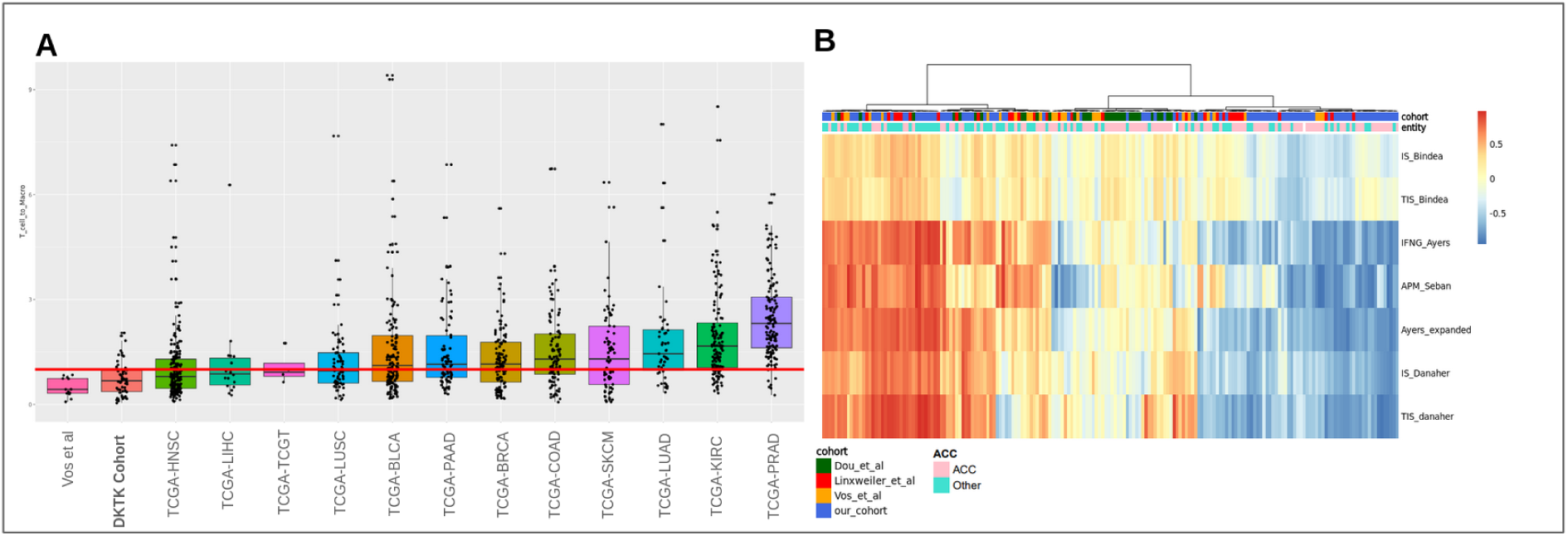
**A:** T-cell to Macrophage ratio from deconvoluted immune cell proportion in a selected set of TCGA cohorts, DKTK SGC cohort and Vos et al SGC cohort. Samples from TCGA were filtered by tumor stage (TNM > 3). Only significant results are displayed **B:**Combined analysis of immune infiltration in multiple cohorts (Vos et al ^9^, Linxweiler et al ^13^, Dou et al ^14^, our cohort). Samples are annotated by entity and cohort.

**Figure S3.**
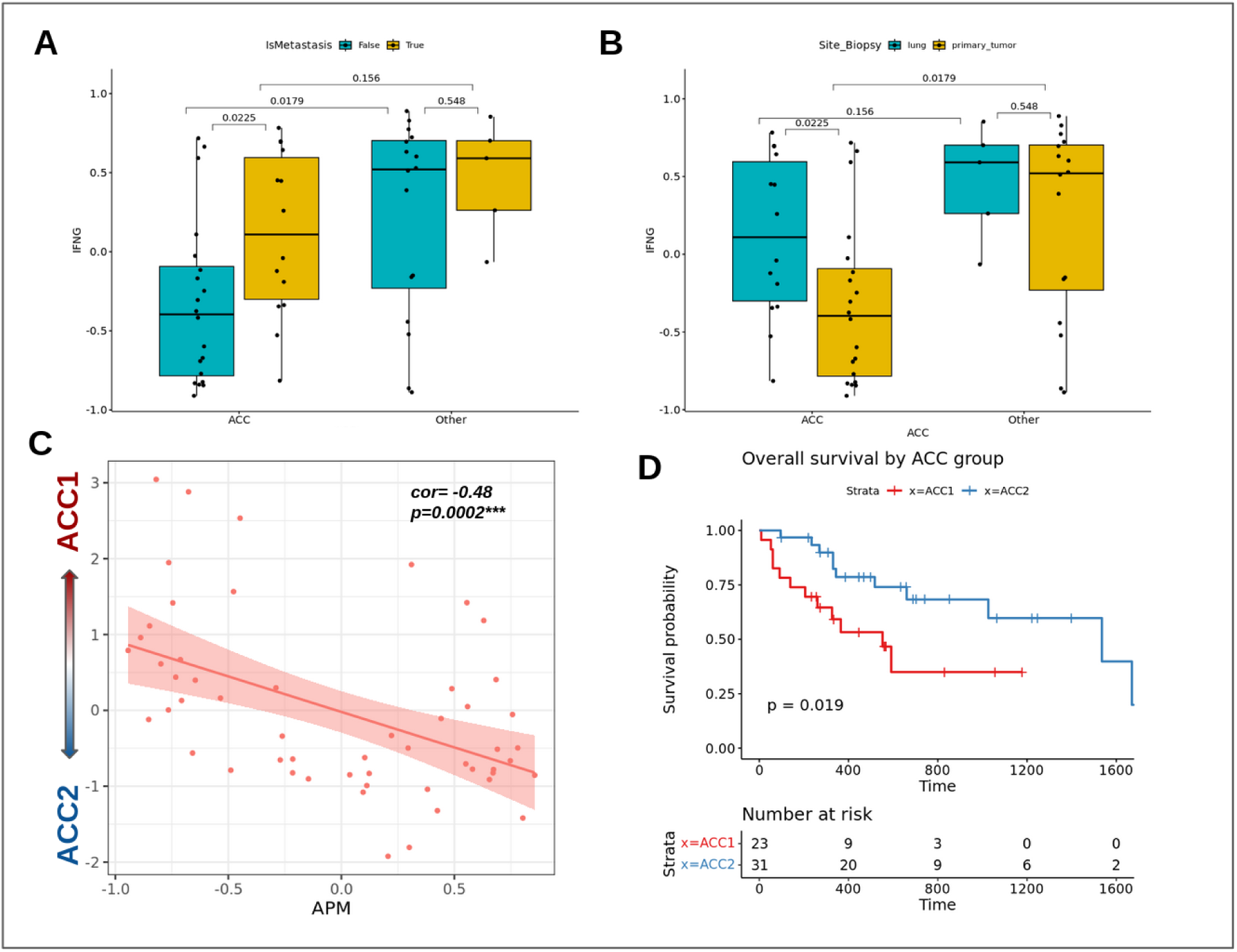
**A:** Association of IFNG score with sample origin (metastatic vs primary) in ACC and non-ACC. **B:** Similar to A only showing the comparison between lung metastases and primary tumors. **C:** Correlation plot between ACC-score and APM score. **D:** Survival plot of ACC1 and ACC2 group.

**Figure S4.**
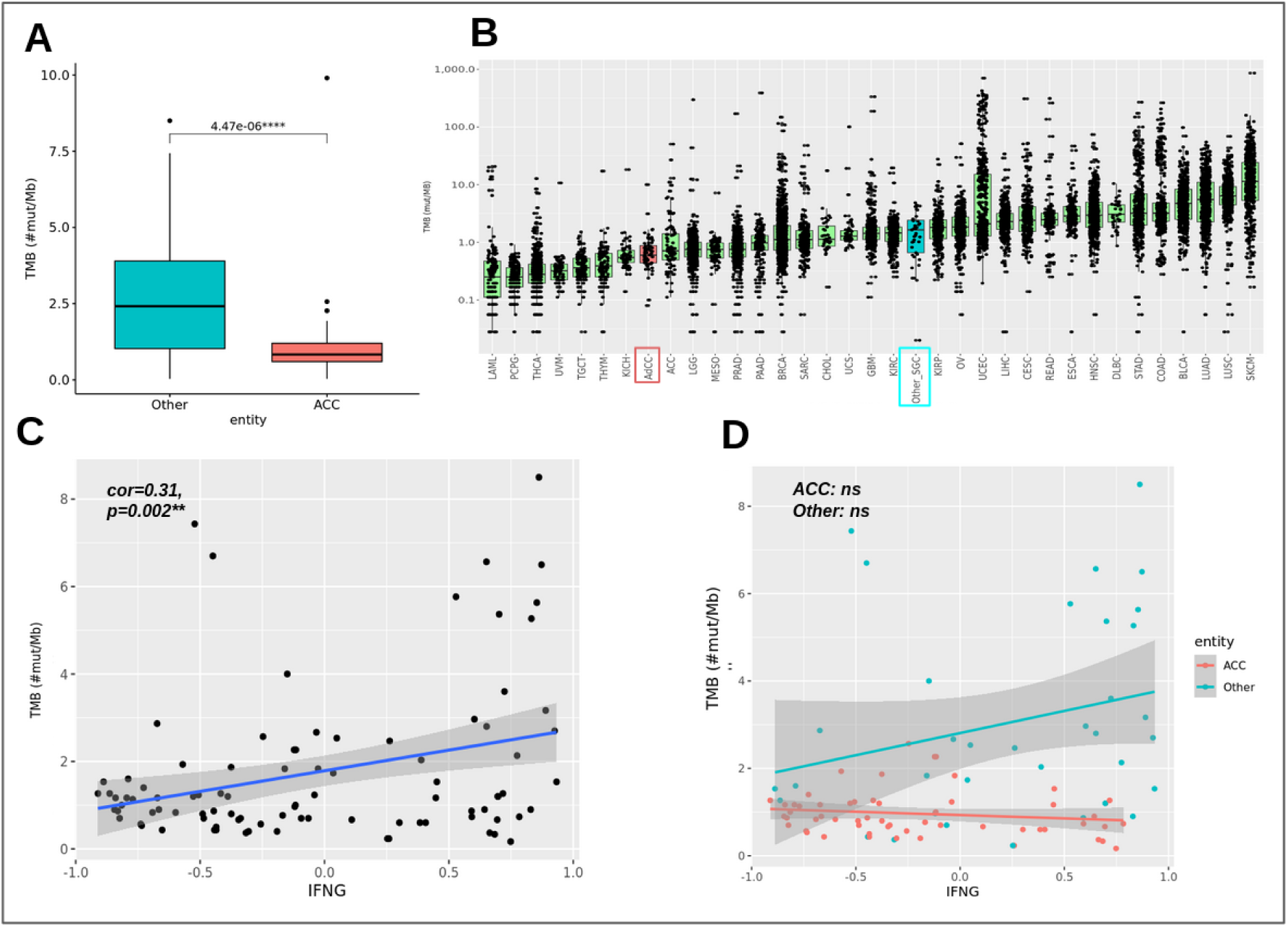
**A:** TMB comparison between ACC (n=62) and other SGC (n=43) entities. **B:** TMB of our cohort (ACC and non-ACC) compared to 33 TCGA cohorts. **C and D:** Correlation plot of IFNG vs TMB of all samples (C) and colored by entity (D).

**Figure S5.**
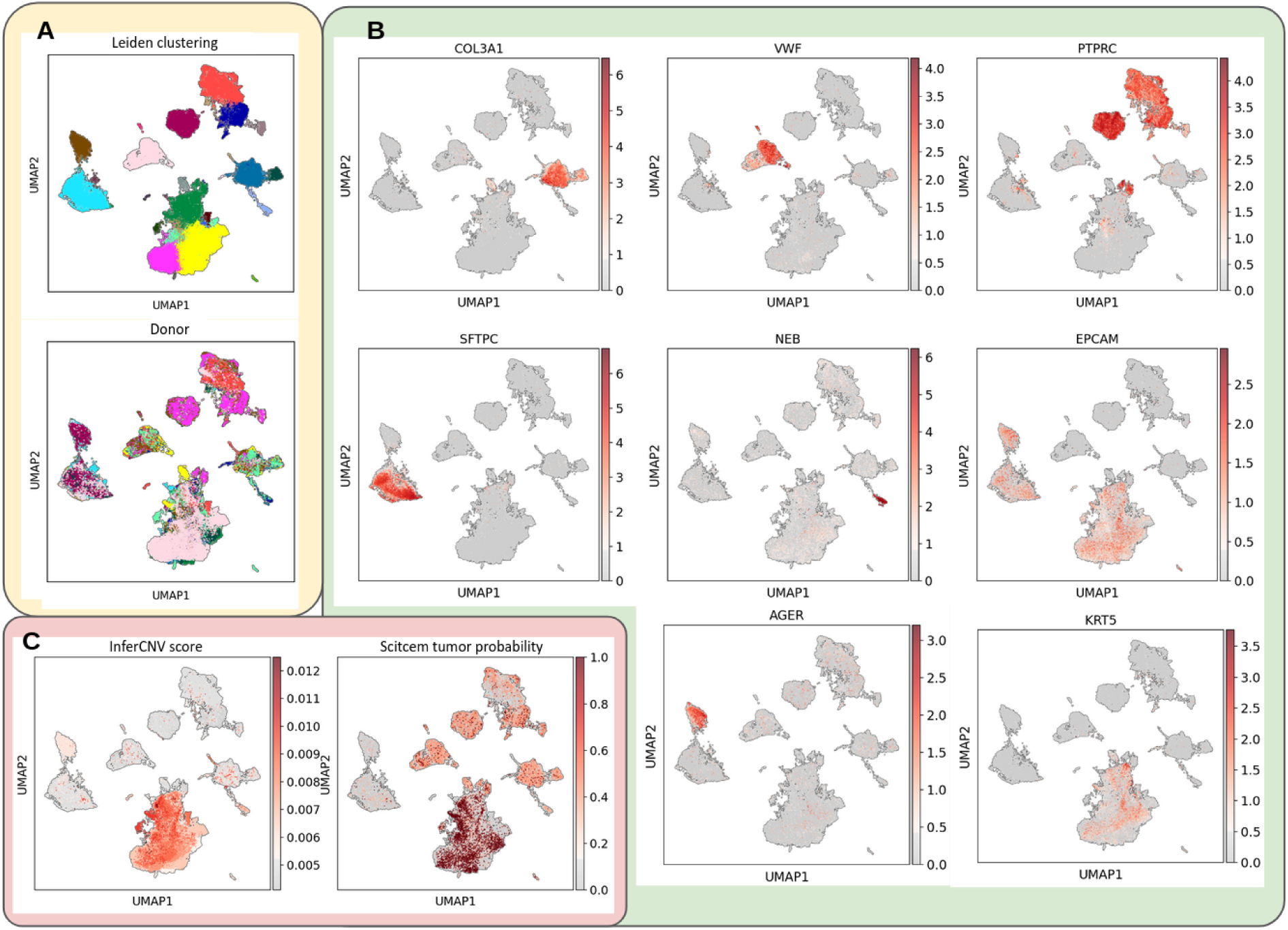
**A:** UMAP of integrated single nuclei data colored by cluster and by patient. **B:** UMAPS with expression of several cell markers (*COL3A1* - Fibroblasts, *VWF* - Endothelial, *PTPRC* - Immune cells, *SFTPC* - Alveolar cells type 2, *AGER* - Alveolar cells type 1, *KRT5* - Basal cells, *EPCAM* - Epithelial cells, *NEB* - Skeletal muscle cells) **C:** UMAPS colored by tumor cell probability as calculated by Scitcem and by InferCNV score.

**Figure S6.**
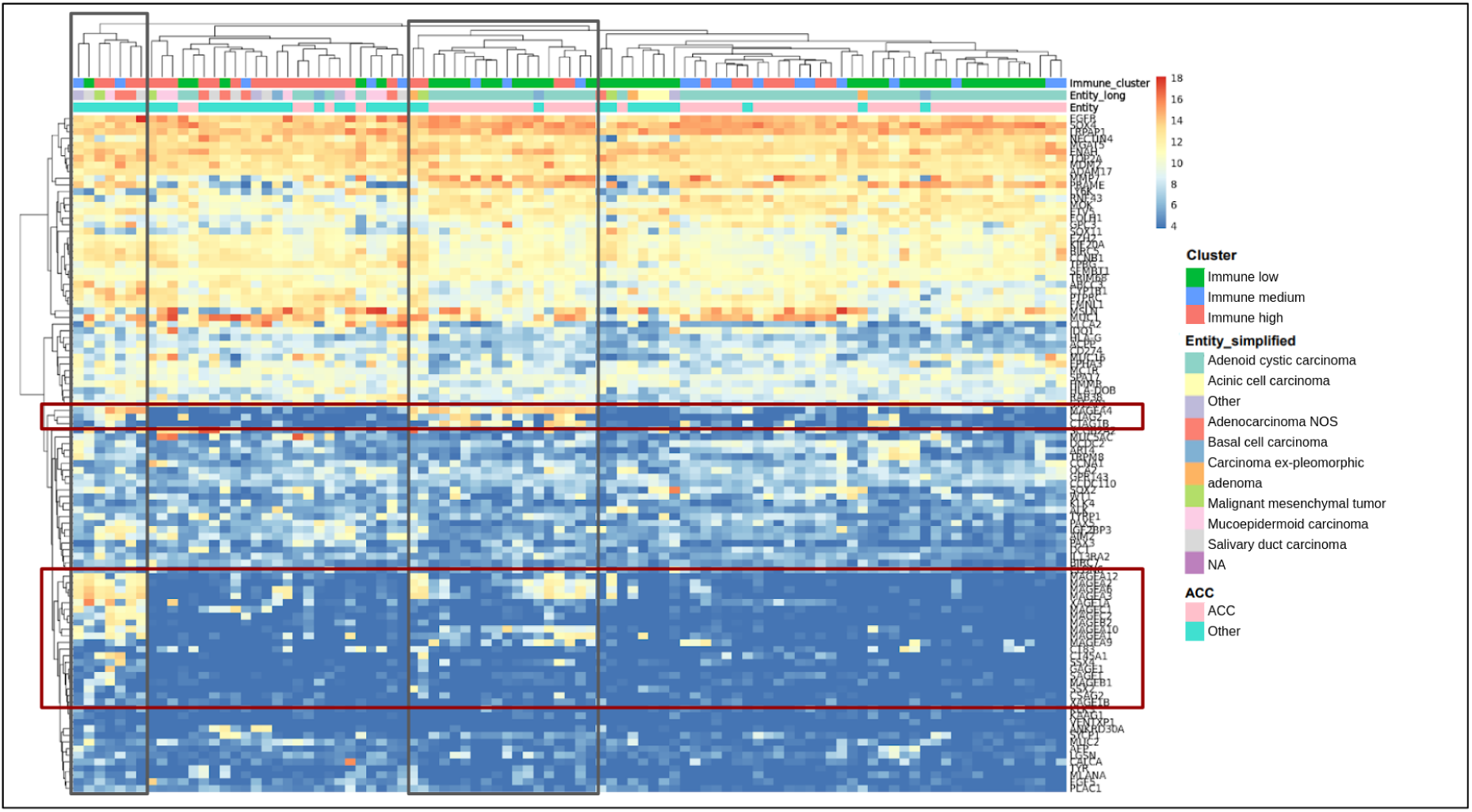
Expression of T-cell-target genes. Samples and genes are clustered. Samples are annotated by Entity and Immune clusters. Marked in red are testis antigens of the MAGE family, marked in black the samples expressing these genes or some of these genes.

## Notes

### Author Declarations

The DKTK MASTER program was approved by ethics committees at all participating sites (Lead Heidelberg, S-206/2011). Written informed consent was obtained from all participants. Ethics approval for retrospective analysis of tumor samples from patients not participating in the DKTK MASTER program was obtained separately (Berlin, EA1/305/21).

### Summary of Updates

- The abstract was extended - The list of authors was updated

